# Analysis of geo-temporal evolution and modeling of the COVID-19 epidemic in Libya

**DOI:** 10.1101/2020.09.19.20197822

**Authors:** Amin Bredan, Hani Benamer, Omran Bakoush

## Abstract

Having been experiencing years of political fragmentation and military conflict, Libya was poorly prepared to meet the challenges of the COVId-19 pandemic. Nevertheless, restriction of travel and social distancing rules were put in place days before the first case was detected. During the next two months, the number of cases grew gradually to 77 cases, followed by a rapid spread that has produced 3691 confirmed cases and 80 deaths by the end of July 2020. The turning point on 26 May 2020 was preceded three weeks earlier by the arrival of the first of a series of flights repatriating Libyans who became stranded abroad when air travel was suspended. In the first weeks of the surge, the number of cases was particularly high in the less densely populated southern region, raising questions about the implementation of social distancing and other protective measures in that region. The epidemic in Libya was modeled using the classical Susceptible-Exposed-Infected-Recovered (SEIR) mathematical model of infectious disease epidemics. Three scenarios were developed based on three estimates of the fraction of the population exposed to the disease (1.5, 2.5 and 3.5%). The modeling portrays the peak of the epidemic around early August and estimates that the number of deaths will flatten out around early November at between 250 and 600, depending on the parameter employed. More deaths than those estimated implies that it is more widespread than assumed. Greater promotion of awareness and understanding of social distancing practices and their value is needed, particularly in the south, and better protection of the elderly should decrease mortality.

---

Libya is a North African country bordering the Mediterranean Sea. It has an area over two and a half times that of France but with an estimated population of only 6.8 million (WordBank 2020). Since the armed conflict that overthrew the regime of Muammar Gaddafi in 2011, Libya has been embroiled in various armed conflicts in various parts of the country. There are two competing governments, one based in the eastern region and the other in the western region. The fractured country has been further crippled by the drop in oil prices compounded by periodic suspension of oil exports. The country was ill prepared to meet the challenges of the COVID-19 pandemic. Despite that, before identification of the first COVID-19 case in Libya, air travel was suspended, schools and universities were closed, and other preventive measures were taken starting on 16 March 2020. Eight days later, the first case of COVID-19 was identified: an elderly man returning from Saudi Arabia *via* Tunisia.

The National Center for Disease Control in Libya (NCDC), which has branches in many cities and has managed to work on a national level, was tasked with overseeing the national effort to combat the COVID-19 pandemic. It publishes information about infections, deaths, recoveries and other information on its Facebook page daily (https://www.facebook.com/pg/NCDC.LY/posts/), but it does not provide the information in tabular or other form that is easily accessible. The data were collected by scrolling its Facebook page, reading all the bulletins, and recording the data from 24/3/2020 until 30/7/2020. The data were entered and analyzed in Excel.

## Analysis of temporal and geographic spread of COVID-19 in Libya

Two months after identification of the first case, the number of COVID-19 cases confirmed by PCR stood at 77 on 26 May 2020, with 3 deaths. The latter date was a turning point after which the number of infections and deaths rose rapidly (Figure 1). By 31 July, there were 3691 confirmed cases, with 80 deaths. This change seems to be associated with two factors: the arrival of Libyan citizens who had been stranded abroad since the suspension of air flights, and the challenges of practicing social distancing in a country abiding by the Arab sociocultural communication norms.

**Figure 1:**
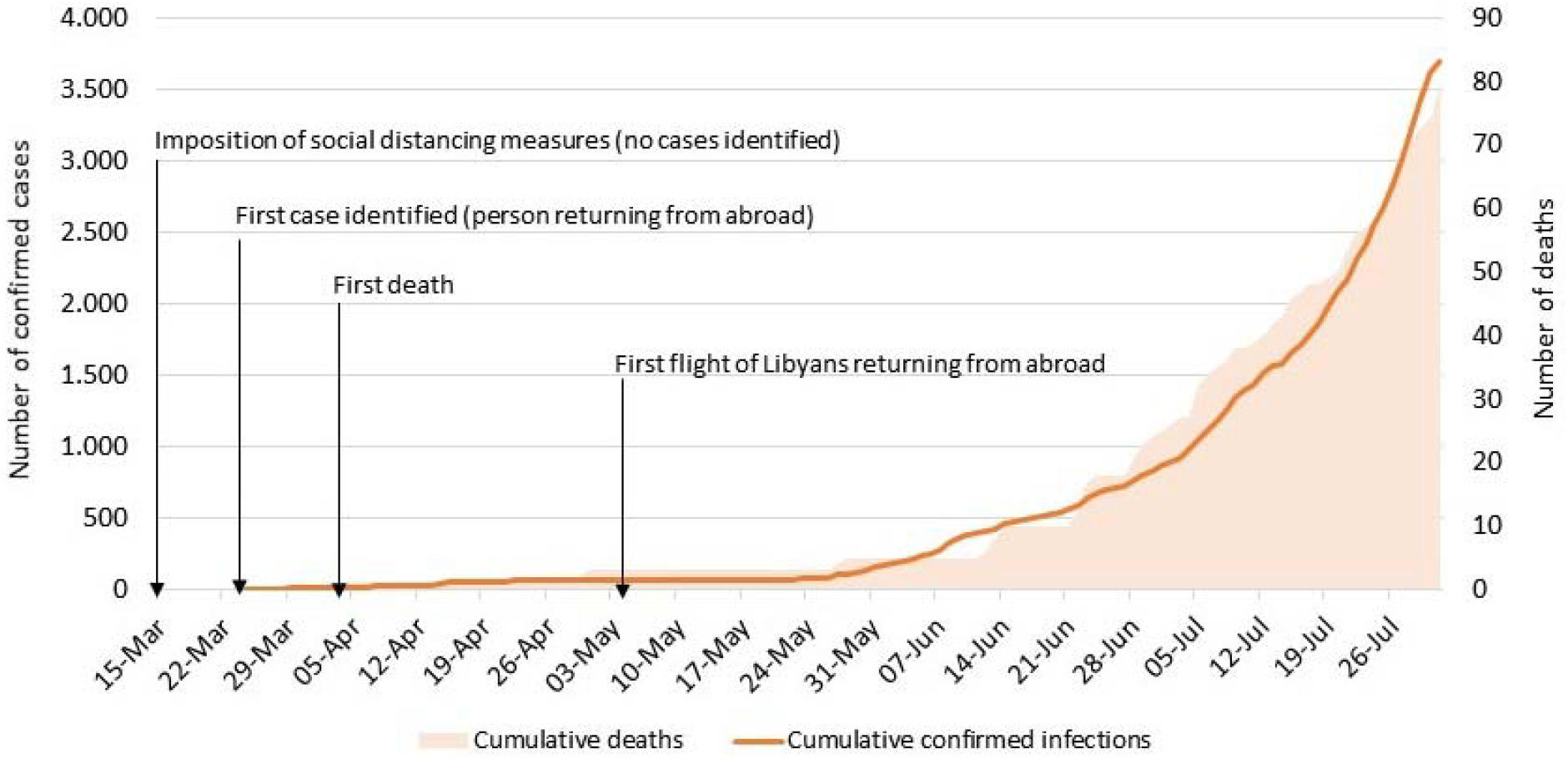
Cumulative numbers of confirmed infections, deaths and landmarks

On May 5^th^, flights chartered to bring home Libyans stranded in North African, Middle Eastern and European countries started to arrive (Figure 1). The Libyan government had made arrangements to have these travelers quarantined and tested by PCR in the countries of their residence at that time before being allowed to board flights to Libya. Unfortunately, the NCDC bulletins show that by 12 July there were 63 confirmed cases of COVID-19 among travelers returning from abroad. Some of these patients may have been infected sometime between having swabs taken for PCR and their arrival in Libya, but there are some unconfirmed reports of some travelers avoiding the quarantine and/or the screening procedures arranged by the Libyan government.

At least initially, some of the travelers were not quarantined upon arrival due to inadequacy of resources. It seems that quarantine measures were better implemented in the eastern region, and much fewer infections have been observed in the eastern region compared to the western and southern regions.

The return of infected Libyan citizens to Libya seems to have been the original source of the infection spread. During the 14 days preceding the start of the rise in infections on 26 May, a total of 11 new confirmed cases were reported nationwide, 9 of whom were Libyans who had returned from abroad. Over the next two weeks, 257 new infections were reported nationwide at 12 locations (Figure 2A) and during the following two weeks there were 494 new cases at over 30 locations (Figure 2B).

**Figure 2:**
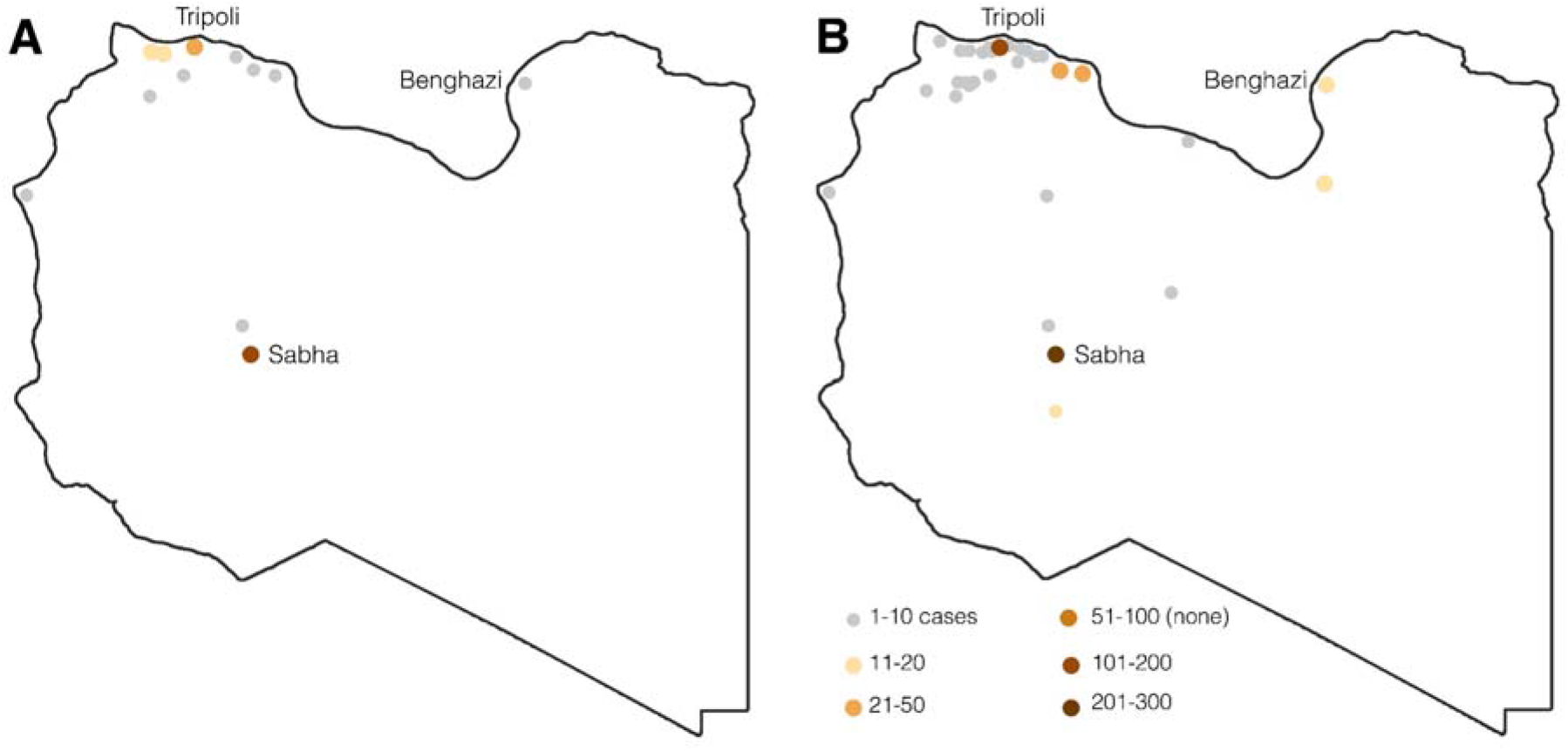
Geographic distribution of laboratory diagnosed cases of COVID-19. (A) 26 May to 8 June. (B) 23 June 6 July

The known imported cases imply a substantially larger number of infected people returning to Libya, either asymptomatic or with mild symptoms that did not motivate them to seek medical attention. Considering that measures such as school closure and partial lockdown or intermittent curfew were introduced even before the start of the epidemic in Libya, in our opinion the protective measures of quarantine and self-isolation were not implemented effectively enough to contain the relatively few known imported cases. Nevertheless, one should not ignore the possibility that the population felt confident that none of the people returning from abroad were infected and interacted with them accordingly.

Based on the assumption that the surge had its origin in the imported cases, one can expect that the spread of the virus would have been more intense in the larger population centers if social distancing and other protective measures were implemented uniformly throughout the country. However, analysis of the geographic distribution of the infections during successive two-week periods, starting with the start of the surge, showed otherwise. Sabha, which has a population approximately one-tenth of that of the capital, Tripoli, had almost threefold more new infections (134 *vs*. 50) during the first two weeks of the surge (Figure 2A). This pattern continued into the subsequent four weeks. The pattern during weeks 5-6 is shown in figure 2B. During weeks 7-8 of the surge, new infections occurred in 45 locations, with Tripoli taking the lead in the number of new infections (297 confirmed cases) while Sabha fell into second place (228 cases).

While it is reasonable to assume that the imported cases of the disease served as the starting point for the disease, it is unlikely that people from Sabha were over-represented among the imported cases to a degree that would explain the large surge in Sabha. More likely, the surge was permitted by the difficulty of social distancing in the local sociocultural environment. In contrast, the protective measures of quarantine and self-isolation for returning travelers may have been more strongly implemented in the eastern region.

## Forecasting the COVID-19 pandemic in Libya

In principle, death is a clear and countable final outcome. Nevertheless, even this clarity is overshadowed by the different definitions of death from COVID-19, and particularly the question of whether to count people who die with symptoms of the disease but no laboratory diagnosis. This situation might exist particularly in Libya and other Muslim countries, where prompt burial is a priority. The COVID-19 case-fatality rate has been reported to be 0.1% (95% CI 0.1-0.2%) for the age group below 65 years and 4.3% (95% CI: 2.7-7.7%) for those of older than 65 years (Riou, Hauser et al. ; Folkhälsomyndigheten 2020). Based on the Libyan population age structure, where only 4% of the population is aged above 65 years (WordBank 2020), the COVID 19 infection fatality rate in Libya is estimated at about 0.3% (95% CI: 0.2% - 0.5%).

To forecast the COVID-19 epidemic Libya, we used the classical Susceptible-Exposed-Infected-Recovered mathematical model of an infectious disease epidemic (Wu, Leung et al. 2020). We fitted the model to the estimated infection fatality rate of 0.3% for the Libyan population, the COVID-19 basic reproduction number (R0) of 2.2, and a duration of infectiousness of 5 days (ECDC 2020; Li, Guan et al. 2020). Then, we estimated the epidemic of COVID-19 among the Libyan population based on the number of deaths due to COVID-19 reported by NCDC up to the end of July 2020. Next, the number of active cases and the number of deaths over the next four months were simulated using the epidemic online calculator (https://gabgoh.github.io/COVID/).

## Spread of COVID-19 in Libya

Based on the trend of the reported deaths in Libya, it is estimated that 1.5% to 2.5% of the Libyan population (*i*.*e*., 100,000 to 170,000 individuals) will have become exposed to COVID-19 by the end of 2020. The estimated number of active cases over time is shown in figure 3. Based on the regional clusters of the reported COVID-19 deaths, the model estimates that 15% of the populations of Sabha, Murzuq and Jufra (100,000–110,000 individuals) will have been exposed to COVID-19 by December 2020.

**Figure 3:**
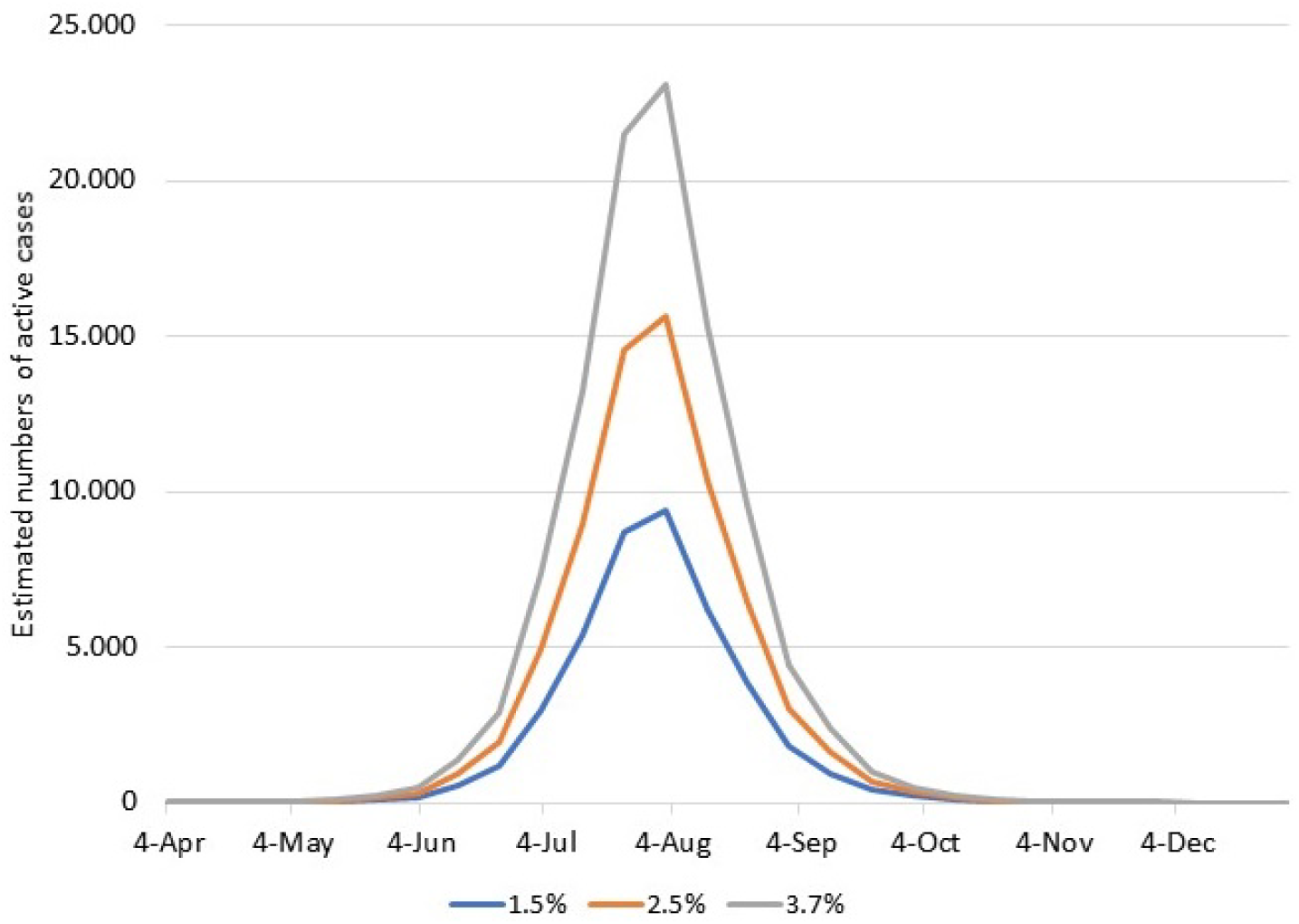
The estimated number of active cases over time if the current spread is constrained to the predicted prevalence of 1.5, 2.5 or 3.7% of the Libyan population.

The Libyan healthcare services are strained and the ability to trace and diagnose all COVID-19 cases could be hampered, which would lead to underreporting of deaths. By December 2020, the cumulative number of deaths is estimated to be in the range of 250-410 (Figure 4). If we assume that COVID-19 deaths are under-reported by 50%, as many as 3.7% of the Libyan population (250,000 individuals) could be exposed to COVID-19 by December 2020. This higher estimate results in a cumulative death estimate of up to 610 cases by December 2020.

**Figure 4:**
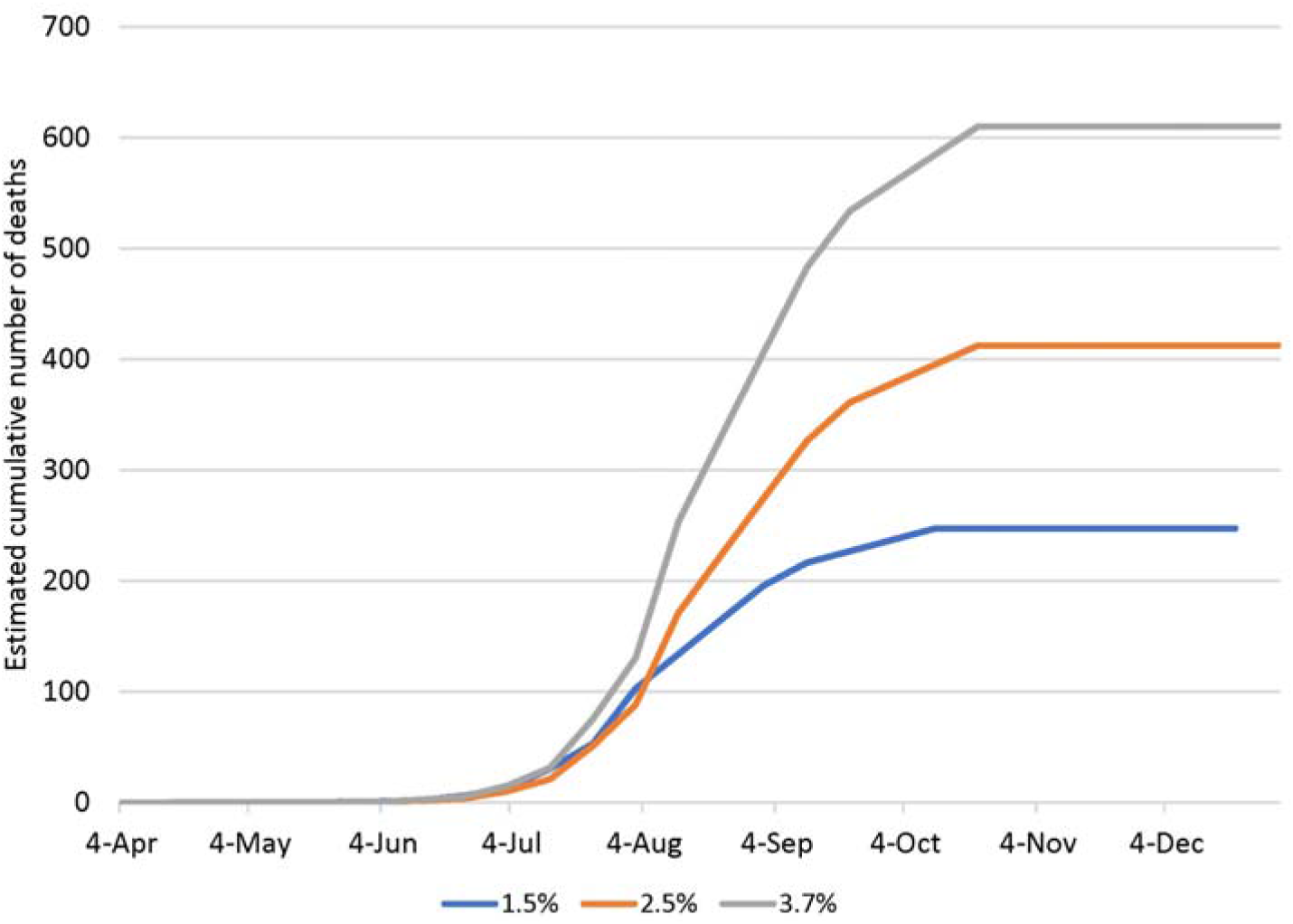
The estimated cumulative number of deaths by December 2020 if the current spread is constrained to the predicted prevalence of 1.5, 2.5 or 3.7% of the Libyan population.

## Recommendation

The protective measures currently applied in Libya could be judged as successful if the number of COVID-19 infections does not exceed 1.5-2.5% of the Libyan population. Of course, the curve can be flattened by better compliance with social distancing rules and implementation of other COVID-19 protection measures. However, if the death rate exceeds the rate projected here, it is a signal that there is a wider spread of the COVID-19 epidemic in Libya. Community transmission in the southern region tends to be large compared to the northern and eastern regions. Thus, emphasizing social distancing measures to slow the spread should be a priority in this region. More importantly, self-isolation of the elderly population is of paramount importance to decreasing the mortality risk.

## Data Availability

The data are available from the website of the Libyan National Center for Disease Control
https://ncdc.org.ly/Ar/

## Notes

**Conflict of interest** The author declares no conflict of interest.

### Competing Interest Statement

The authors have declared no competing interest.

### Funding Statement

No funding was received for this work.

### Author Declarations

All the data used in this study have been made publicly available by the Libyan National Center for Disease control on its website, and no approval was needed.

